# Modified Harvard Step Testing within a Clinic Setting Enables Exercise Prescription for Cancer Survivors

**DOI:** 10.1101/2020.09.30.20204776

**Authors:** Katherine R. White, Jana Lu, Zara Ibrahim, Priscilla A. Furth

## Abstract

**Purpose:** To evaluate the utility of modified Harvard Step tests within the context of a comprehensive physical examination for fitness evaluation and exercise prescription for cancer survivors.

**Methods:** A retrospective chart review of initial cancer survivor clinic visits over a ten-year period (n=169) was conducted to evaluate correlations between demographic factors, clinical characteristics, step and strength test performance, and exercise prescriptions.

**Results:** Clinic population was 94% female, aged 27-79 years, predominantly breast cancer (87%), presenting within two years of cancer diagnosis with current exercise history significantly less vigorous than past exercise (*p*=0.00; 34% sedentary). Fifty-two percent completed a 3-minute-9-inch step test at pace (96 steps per minute). Fourteen percent required slower self-pacing, 12% both a slower pace and shortened time, and 5% a flat test. Younger age (*p*=0.04) and more vigorous exercise histories (*p*<0.04) correlated with ability to complete the at pace test but all formats led to exercise prescriptions more vigorous than current activity (*p*<0.0002). Stratified fitness ratings using YMCA normative data yielded associations between higher fitness levels and lower BMI (F(1,86)=4.149, *p*<0.05), office pulse (F(1,87)=7.677, *p*<0.05), and systolic blood pressure (F(1,18)=6.58, *p*<0.05).

**Conclusions:** Office-based fitness evaluation with a panel of modified step test options accommodating different baseline fitness levels enabled personalized exercise prescriptions more vigorous than current activity.

**Implications for Cancer Survivors:** Cancer patients frequently engage in less vigorous activity as they enter into survivorship. Modified step tests are a means for office-based evaluation of cardiovascular fitness within the context of a comprehensive physical examination.

## BACKGROUND

Cancer survivors are a rapidly increasing population globally due to improved screening, diagnoses, and treatments. In the United States alone, there are more than 15.5 million cancer survivors; this number is expected to double in a few decades [1]. For years, clinicians advised cancer patients to refrain from physical activity; however, in the late 1990s, research outlined the benefits of physical activity, including improving survivorship, physical well-being, and quality of life [1–3]. The American Cancer Society (ACS) and the American College of Sports Medicine (ACSM) recommend cancer survivors engage in at least 150 minutes of moderate or 75 minutes of vigorous aerobic activity per week, with biweekly resistance training and daily muscle stretching [1,3]. At a minimum, current guidelines urge survivors to avoid inactivity. In 2018, the American Heart Association emphasized the importance of integrating physical activity assessment in routine clinical practices and workflow [4]. Recent studies have found that cancer survivors who exercise have lower relative risks of cancer mortality, improved quality of life, and reduced fatigue [5–8]. Despite these recommendations, up to 80 percent of cancer survivors may not meet ACS and ACSM physical activity recommendations and both survivors and providers are not always aware of the all the recommended exercises, which include aerobic, balance, stretching, and muscle and bone strengthening [2,3,9].

Cancer survivors face unique cardiovascular fitness challenges. Chemotherapy and radiation therapy may cause cardiotoxicity, potentially impairing exercise ability [10]. Patients with hormone receptor-positive operable breast cancer treated with chemo-endocrine therapy have lower peak exercise stroke volume, cardiac output, cardiac power output and reserve, and VO2peak than their healthy counterparts [11]. Exercise interventions have been shown to increase fitness, physical function, and quality of life in prostate cancer survivors treated with hormone therapy [12,13]. Overweight or obese survivors experience increased cancer risk and decreased overall survival [14]. Hypertension also increases risk of cancer mortality [15]. Although cancer patients with distant metastasis are not typically counseled on exercise, studies have found that exercise in advanced cancer can improve fitness, function, fatigue, and overall quality of life [16,17].

It is important for cancer survivors to consult a physician before engaging in physical fitness programs, to individualize physical activity to current level of physical conditioning and personal preference [1,3,18,19]. While exercise is possible to initiate at all ages, careful attention must be given for older individuals who may be at a greater risk of falls, injuries, sudden cardiac death or myocardial infarction [3,20]. There are limited numbers of peer-reviewed studies addressing how primary care providers may provide exercise prescriptions, especially for cancer survivors. Many times this is left to the oncologist. Formal training in physical fitness assessment is frequently lacking in medical school curriculum and may be limited to specific medical specialties such as physical medicine and rehabilitation [21–23].

Approaches for fitness assessment in the office setting include self-report or direct fitness testing [24]. Comprehensive fitness assessment should assign intensity to past and current exercise history, cardiovascular fitness, muscle strength, balance, and flexibility [25]. Self-reported physical activity can be coded with respective Metabolic Equivalency of Task (MET), a reflection of required oxygen uptake [26]. Exercise intensity can also be graded according to VO2max percentages with low intensity classified as <37% VO2max or between 37-45% VO2max, moderate intensity as 46-63% VO2max, and high intensity as 64-90% VO2max or ≥91% VO2max [18].

The Harvard Step Test is one tool for fitness assessment. Since the initial development of the Harvard Step Test for evaluation of military recruits in World War II, the basic technique has been evaluated with different modifications and successfully applied across different settings [27]. The YMCA has developed age and gender-adjusted standards for fitness rating using one minute pulse recovery standard for a 12-inch-3-minute-96 steps/minute format [28]. Reported correlates of fitness in adults include positive associations with male sex, education, socioeconomic status, and leisure-time physical activity and inverse relationships with age, body mass index (BMI) and resting heart rate [29]. Increased cardiorespiratory fitness is associated with lower hypertension risk [30].

Our study evaluates modifications of the Harvard Step Test for fitness evaluation in cancer survivors in the context of a comprehensive physical examination. For our population, the step test height was reduced to 9 inches, and survivors were afforded the option of self-pacing at a slower pace and/or stopping the test prior to three minutes. Previous research has shown that modifications using an 8 inch step test with a limit of two minutes were an effective substitute for the six minute walk test, another common measure of exercise capacity [31]. A step test requires more exertion than a walk test, and has been reported as a better choice for patients with higher baseline fitness levels [32]. For those with lower baseline fitness levels we utilized a 3-minute step-in-place protocol (flat test) that could be performed inside the examination room. Cancer survivors seen in our clinic were also tested for range of motion, muscle strength, tone, rigidity, posture, and gait as part of their neurological exam. We also explored the use of submaximal (max) muscle strength testing: a 20-max-crunch test for evaluation of abdominal muscle endurance, 20-max-modified-push-ups from the knees for evaluation of overall muscle strength and endurance, a 5-max-squat test for evaluation lower extremity muscle weakness and tightness, and a 20-second-max plank (prone bridge) for core strength [33–35]. Both step and submaximal muscle testing was incorporated into a comprehensive examination that included taking specific past and current exercise histories.

## METHODS

We conducted a retrospective chart review of all patients presenting to the Lombardi Fitness and Metabolism Clinic between 2008 and 2018. This study was approved by the Georgetown University Institutional Review Board and determined to impose minimal risk on participants. A waiver of consent was obtained.

### Eligibility

In order to be included in the study, patients needed to be cancer survivors presenting for their first visit between 2008 and 2018. One hundred and sixty-nine of 171 patients (99%) met the selection criteria. Two patients seen in the clinic during this time were excluded from analysis as not having been diagnosed with cancer (n=1 Prophylactic bilateral mastectomy for elevated breast cancer risk; n=1 Chronic pulmonary disease.).

### Data Collection

Detailed chart reviews in the Aria and Powerchart Electronic Medical Records were conducted to ascertain demographic factors (sex, age, employment status, occupation, residence, living situation), clinical characteristics (cancer type, pathologic stage, site of metastases, age at diagnosis, treatment history, BMI, resting pulse, systolic blood pressure), and fitness measures (self-reported past and current exercise histories, step test completion, pulse recovery, core testing, exercise prescription) at the time of initial office visit. All patients underwent a comprehensive history and physical (H&P) prior to office fitness evaluation. Occupations were categorized by economic sector. Cancer types were classified into five organ systems (Breast; Gastrointestinal (GI) including Pancreatic, Gastric, Ampulla of Vater, Rectal; Head and Neck including Tongue, Tonsil, Merkel Cell; Genitourinary (GU) including Ovary, Prostate, Renal Cell; Hematopoietic including Lymphoma, Angiosarcoma, Multiple Myeloma). Presence of distant metastasis was defined as spread from primary tumor to distant organs or distant lymph nodes. Surgical procedures related to diagnosis and/or treatment of the primary cancer were recorded. Self-reported past and current exercise histories were coded as sedentary, light, moderate, or vigorous [26,36]. Data was stratified by the type of submaximal cardiovascular fitness assessment performed: Completed At Pace (96 +/-0 steps/min, 9 inch) (n=88), Completed Shorter Self-Paced (91+/-1.5 steps/min, 9 inch) (n=24), Completed Shorter Self-Paced (91+/-1.5 steps/min, 9 inch) (n=21), Flat Self-Paced (94+/-1.6 steps/min) (n=9) or whether it was contraindicated (n=19) or deferred (n=8). Step testing was contraindicated by H&P for active pain, severe deconditioning, indications for cardiac stress test, onycholysis, wound dehiscence, balance impairment. Step testing was deferred for non-fitness chief complaints or when patient records were adequate to assess cardiovascular fitness. One patient was deferred due to a need to leave their appointment early. For those that completed a 9-inch step test, age and 1-minute pulse recovery were used to assign YMCA Fitness Category for patients who completed step testing (Very Poor-Poor, Below Average, Average, Above Average, Good, Excellent) [28]. Results of core strength testing (numbers of crunches and/or modified push-ups from the knees, squat testing (normal defined as symmetric performance of five serial squats) and plank duration were recorded and presented for each category of cardiovascular fitness assessment including those for which a step test was contraindicated or deferred [34].

### Statistical Analyses

Mean, standard deviation, and range were calculated for years since diagnosis, age at clinic presentation, age at diagnosis, BMI, initial office pulse, and initial office systolic blood pressure (GraphPad Prism version 8.4.3for macOS, GraphPad Software, San Diego, California USA, www.graphpad.com). Associations between categorical variables were analyzed using Chi Square (https://www.socscistatistics.com/tests/chisquare2/default2.aspx) and Fisher’s Exact (http://vassarstats.net/fisher2x4.html; https://astatsa.com/FisherTest/) tests (accessed July-September 2020). One-way ANOVA followed by Tukey’s multiple comparisons test was performed to examine associations between continuous and categorical variables (GraphPad Prism). Associations between two continuous variables were analyzed using linear regression (GraphPad QuickCalcs website, https://www.graphpad.com/quickcalcs/linear1/ (accessed July 2020). *P*<0.05 was considered statistically significant.

## RESULTS

### Most Cancer Survivors Presenting to Clinic Were Actively Employed with Established Places of Residence

The majority of cancer survivors were actively employed at the time of presentation (76%), largely within the Education, Research and Development (21%) and Service economic sectors (60%) (Table 1). Individuals uniformly held established places of residence with living situations divided across living with a life partner (34%), living with family including children (32%), and living alone (29%). For some individuals information was not available in the chart (10% for employment status, 19% for employment, 4% for living situation).

**Table 1:** Living, Employment, and Occupations of Clinic Participants.

|  |  |
| --- | --- |
| **Clinic Total** | **171** |
| **Cancer Survivor Total, No. (%)** | **169 (100)** |
| **Employment Status, No. (%)** |  |
| Currently working | 128 (76) |
| Not currently working | 23 (14) |
| Information unavailable | 18 (10) |
| **Economic Sector, No. (%)** |  |
| ***Service*** | **102 (60)** |
| Business/Finance/Consultant | 20 |
| Art/Design/Media/Journalism | 16 |
| Administration/HR/Manager/Director | 20 |
| Healthcare | 19 |
| Public/Government Service | 15 |
| Law | 12 |
| ***Education, Research and Development*** | **35 (21)** |
| Education/Academia | 21 |
| Science/Technology/Engineering | 14 |
| ***Information unavailable*** | **33 (19)** |
| **Residence, No. (%)** |  |
| Established Place of Residence | 169 (100) |
| **Living Situation, No. (%)** |  |
| Lives only with life partner | 57 (34) |
| Lives with family including children | 55 (32) |
| Lives alone | 49 (29) |
| Lives with roommate | 1 (1) |
| Information unavailable | 7 (4) |

### Different Modifications of the Harvard Step Test Were Used to Assess Cardiovascular Fitness

The majority of cancer survivors presented within two years of diagnosis but there was a wide range from 0 to 26 years (Table 2). The majority were female breast cancer survivors (87%) with the remainder GI, Head and Neck, GU, Hematopoietic and Lung cancer survivors. A minority (9%) had distant metastasis at the time of clinic presentation. The mean BMI was in the overweight range (28 +/- 0.46, mean +/- SEM) with a range from 17-44. Initial office pulse and systolic blood pressures means were in the normal range but the range was wide (56-117 and 94-188, respectively). The majority of survivors presented after completing at least one surgical procedure (93%) and 41% had completed radiation therapy. At the time of presentation 28% were on active chemotherapy and 48% on active anti-hormonal therapy. Only 2% were under active radiation therapy. Sixteen percent of survivors had a life-time history of sedentary activity; however, this proportion increased significantly to 34% at the time of presentation while the proportion of survivors currently engaged in vigorous exercise significantly decreased to 10% from 34% for past exercise history (p< 0.00001, Chi Square, two-tailed). Ninety percent of individuals seen in clinic received an exercise prescription that was significantly more vigorous than current activity for the majority (p=0.0000, Chi Square, two-tailed; p<0.0004, Fisher’s Exact, two-tailed). Exercise prescriptions were deferred for individuals requiring consultative evaluations (n=8 cardiac stress test, n=1 each: exercise-induced cardiac arrythmia, MRI shoulder, Baker’s Cyst, left upper quadrant pain) and further healing (n=1 each: onycholysis, surgical incision). One patient declined an exercise prescription, attending the clinic for consultation on tamoxifen treatment.

**Table 2:**
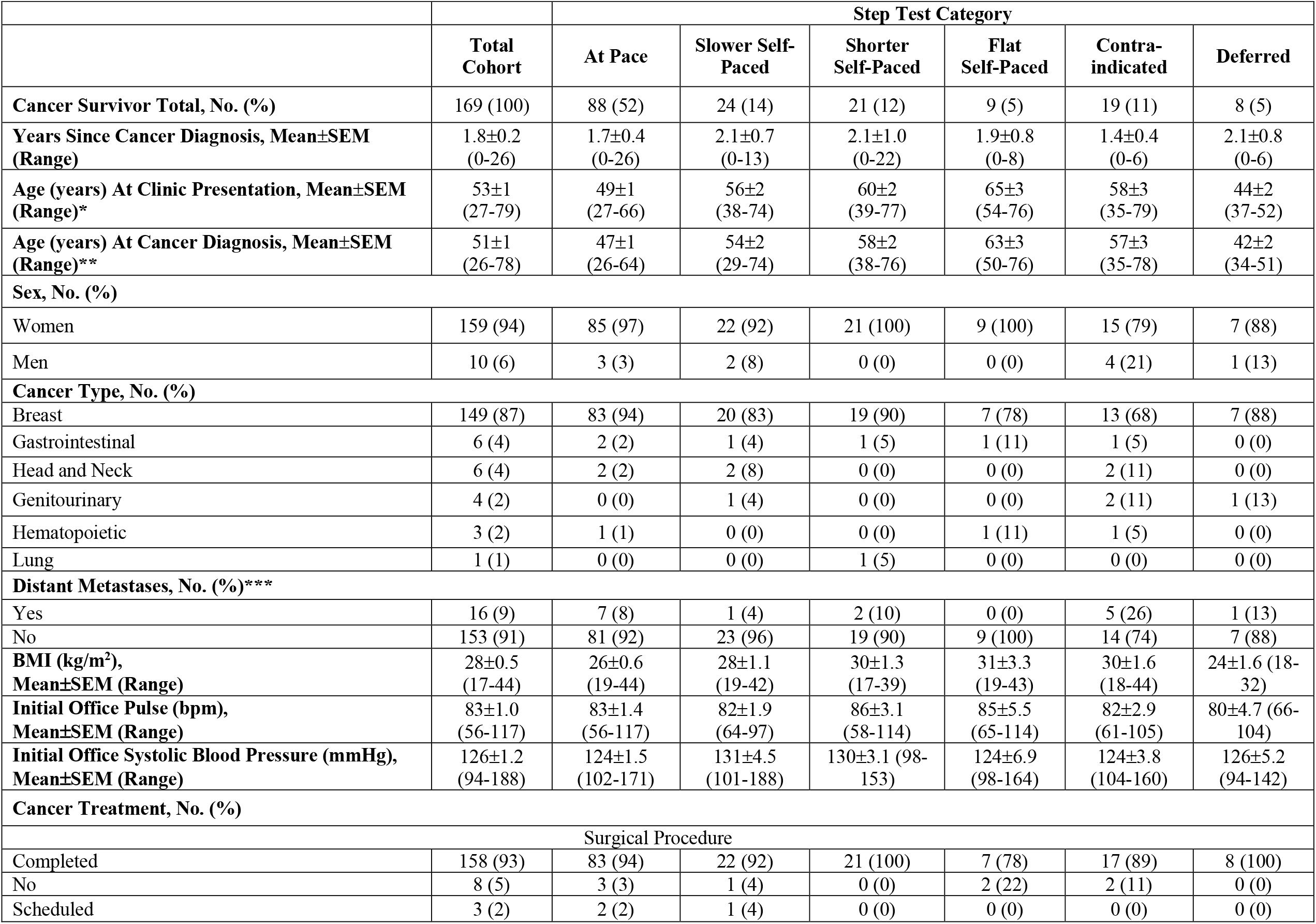
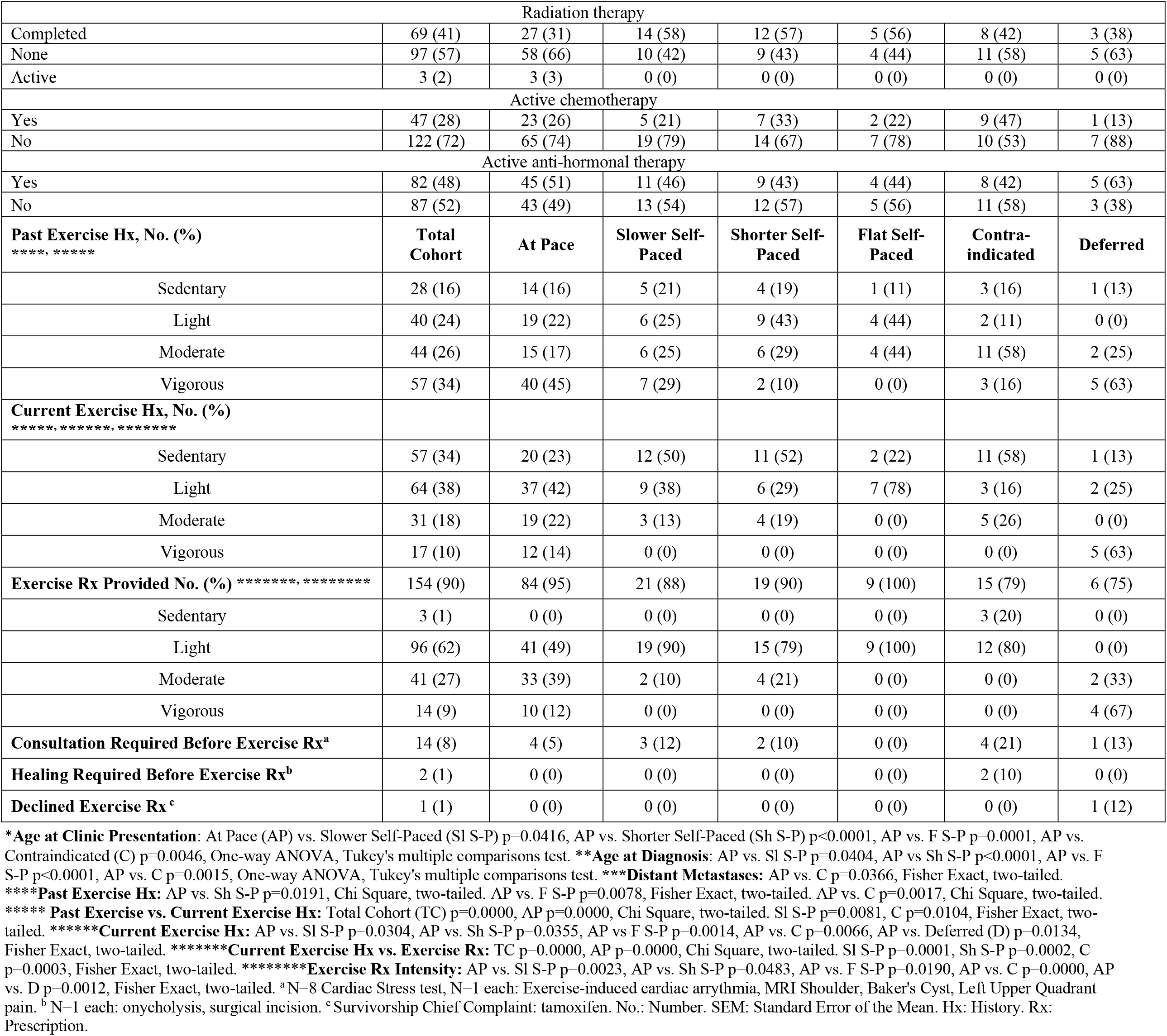
Characteristics, Exercise History, and Prescription by Step Test Category for Cancer Survivors Participating in Clinic.

A one-minute pulse recovery was measured for individuals completing a step test. The largest group (52% of survivors) completed a modified 9-inch, 3-minute, 96 steps/min modified Harvard Step test (At Pace). Step test adjustments were required for the remainder of presenting cancer survivors. Fourteen percent completed the test when allowed to self-modify their pace (85+/-1.1 steps/min, mean +/- SEM) (Slower Self-Paced) as needed due to current poor conditioning/sedentary behavior (n=15), chemotherapy-related fatigue/anemia (n=4), chronic knee/hip pain/arthritis (n=4), or post-chemotherapy neuropathy (n=1). Another 13% attempted a self-paced test (91+/-1.5 steps/min but stopped early (1.5 +/- 0.1 min) (Shorter Self-Paced) for fatigue due to poor conditioning (n=8) or chemotherapy (n=1), shortness of breath (n=4), chronic knee pain (n=3), chemotherapy-related neuropathy pain (n=2), leg prosthesis pain (n=1), leg lymphedema pain (n=1), or discomfort with balance (n=1). Self-paced step testing was limited to a flat surface for 5% of individuals based on non-cancer related conditions identified on H&P (n=7 chronic knee pain; n=2 impaired balance). For 12% of presenting survivors, step testing was contraindicated based on conditions identified on H&P (n=5 active knee pain, n=4 active back pain, n=3 severe deconditioning, n=2 chronic atrial fibrillation, n=2 cardiac stress test indication, n=1 each: impaired balance, onycholysis, wound dehiscence, post-surgical abdominal pain). Step testing was deferred for 5% due to clear documentation of high cardiovascular fitness by history (n=6) or the need to leave appointment early (n=1).

Cancer survivors able to complete the At Pace step test were on average seven to sixteen years younger at the age of clinic presentation and cancer diagnosis (p<0.05, One-way ANOVA, Tukey’s multiple comparisons test) and described more vigorous past and/or current exercise histories (p<0.04, Chi Square, two-tailed or Fisher’s Exact, two-tailed) as compared to individuals from most other groups. The exception was the Deferred group, which was not significantly different by age and showed significantly more vigorous current activity (p<0.005, Fisher’s Exact, two-tailed). The At Pace group also received exercise prescriptions that were significantly more vigorous than other groups (p<0.05, Fisher’s exact, two-tailed) with the exception of the Deferred group who received prescriptions significantly more vigorous than the At Pace group (p=0.002, Fisher’s Exact, two-tailed). There were no statistically significant differences between any groups by sex, cancer type, presence or absence of distant metastases, BMI, initial office pulse or blood pressure, or cancer treatment parameters.

### All Three 9-inch Step Tests Enabled Fitness Stratification But Appeared Most Robust for the At Pace Group Completing the 3-minute Step Test

YMCA age-adjusted fitness ratings were applied to all step test groups. Results from all three groups completing a 9-inch step test could be stratified. The At Pace group stratified from Very Poor to Excellent with the Slower and Shorter Self-Paced groups stratifying from Below Average to Excellent (Table 3). In contrast, results from the group completing a step test on a flat surface showed minimal stratification. To determine if the fitness ratings obtained from the different groups associated with reported fitness correlates, we compared significant associations between the three groups. The number of associations was highest for the At Pace group. Significant regression equations were found for BMI (F(1,86)=4.149, p<0.05), R2 0.046, YMCA Fitness Category decreased 0.061 for each kg/m2) and initial office pulse (F(1,87)=7.677, p<0.05), R2 0.081, YMCA Fitness Category decreased 0.0336 for each beat/min) with a significant positive correlation with active hormone therapy (p= 0.0347, Fisher’s Exact, two-sided) and a negative correlation with active chemotherapy (p=0.0027, Fisher’s Exact, two-sided) (Table 3, Supplementary Figure 1). For the slower self-paced group, there were no associations with reported fitness correlates and active hormone therapy showed a negative correlation (p=0.0328, Fisher’s Exact, two-sided) (Table 3, Supplementary Figure 2). For the Shorter Self-Paced group, significant regression equations were found for initial office systolic blood pressure (F(1,18)=6.58, p<0.05), with an R2 of 0.2676. Participants’ predicted YMCA Fitness Category was equal to 6.237 -0.0164 (initial systolic blood pressure) when blood pressure was measured in mmHg. YMCA fitness category decreased 0.0164 for each mmHg) with a significant positive correlation for active chemotherapy (p=0.0103, Fisher’s Exact, two-sided) (Table 3, Supplementary Figure 3). Taken together, these findings showed while results from all three 9-inch step test groups could be stratified, only the At Pace group completing the 3-minute Step Test showed significant associations with known fitness correlates.

**Table 3.** Distribution of Fitness Ratings by Cancer Treatment and Metastatic Disease Status for Cancer Survivors Completing 9-inch Step Tests.

| **Fitness Ratings** | **Very Poor-Poor** | **Below Average** | **Average** | **Above Average** | **Good** | **Excellent** |
| --- | --- | --- | --- | --- | --- | --- |
| **Completed At Pace (96±0 steps/min, 9 inch), No. (%)<sup>a</sup>** |  |  |  |  |  |  |
| **All** | 4 (5) | 5 (6) | 13 (15) | 18 (21) | 30 (34) | 17 (19) |
| **Cancer Treatment** |  |  |  |  |  |  |
| Radiation therapy |  |  |  |  |  |  |
| Completed | 2 (8) | 0 (0) | 5 (19) | 6 (23) | 8 (31) | 5 (19) |
| None | 2 (3) | 5 (9) | 8 (14) | 10 (17) | 21 (36) | 12 (21) |
| Active | 0 (0) | 0 (0) | 0 (0) | 2 (67) | 1 (33) | 0 (0) |
| Active chemotherapy* |  |  |  |  |  |  |
| Yes | 1 (4) | 3 (13) | 9 (39) | 3 (13) | 4 (17) | 3 (13) |
| No | 3 (5) | 2 (3) | 4 (6) | 15 (23) | 26 (41) | 14 (22) |
| Active anti-hormonal therapy** |  |  |  |  |  |  |
| Yes | 2 (4) | 2 (4) | 2 (4) | 12 (27) | 15 (33) | 12 (27) |
| No | 2 (5) | 3 (7) | 11 (26) | 6 (14) | 15 (36) | 5 (12) |
| **Presence of Distant Metastases** |  |  |  |  |  |  |
| Yes | 0 (0) | 2 (29) | 1 (14) | 2 (29) | 1 (14) | 1 (14) |
| No | 4 (5) | 3 (4) | 12 (15) | 16 (20) | 29 (36) | 16 (20) |
| **Completed Slower Self-Paced (85±1.1 steps/min, 9 inch), No. (%)<sup>b</sup>** |  |  |  |  |  |  |
| **All** | 0 (0) | 1 (4) | 1 (4) | 7 (29) | 9 (38) | 6 (25) |
| **Cancer Treatment** |  |  |  |  |  |  |
| Radiation therapy |  |  |  |  |  |  |
| Completed | 0 (0) | 1 (7) | 0 (0) | 5 (36) | 4 (29) | 4 (29) |
| None | 0 (0) | 0 (0) | 1 (10) | 2 (20) | 5 (50) | 2 (20) |
| Active | 0 (0) | 0 (0) | 0 (0) | 0 (0) | 0 (0) | 0 (0) |
| Active chemotherapy |  |  |  |  |  |  |
| Yes | 0 (0) | 0 (0) | 1 (20) | 1 (20) | 3 (60) | 0 (0) |
| No | 0 (0) | 1 (5) | 0 (0) | 5 (26) | 7 (37) | 6 (32) |
| Active anti-hormonal therapy*** |  |  |  |  |  |  |
| Yes | 0 (0) | 1 (10) | 0 (0) | 5 (45) | 5 (45) | 0 (0) |
| No | 0 (0) | 0 (0) | 1 (8) | 2 (15) | 4 (31) | 6 (46) |
| **Presence of Distant Metastases** |  |  |  |  |  |  |
| Yes | 0 (0) | 0 (0) | 0 (0) | 0 (0) | 1 (100) | 0 (0) |
| No | 0 (0) | 1 (4) | 1 (4) | 7 (30) | 8 (35) | 6 (16) |
| **Completed Shorter Self-Paced (91±1.5 steps/min, 9 inch), No. (%)<sup>c</sup>** |  |  |  |  |  |  |
| **All** | 0 | 3 (15) | 2 (10) | 5 (25) | 6 (30) | 4 (20) |
| **Cancer Treatment** |  |  |  |  |  |  |
| Radiation therapy |  |  |  |  |  |  |
| Completed | 0 (0) | 2 (18) | 2 (18) | 2 (18) | 3 (27) | 2 (18) |
| None | 0 (0) | 1 (11) | 0 (0) | 3 (33) | 3 (33) | 2 (22) |
| Active | 0 (0) | 0 (0) | 0 (0) | 0 (0) | 0 (0) | 0 (0) |
| Active chemotherapy**** |  |  |  |  |  |  |
| Yes | 0 (0) | 0 (0) | 0 (0) | 0 (0) | 5 (71) | 2 (29) |
| No | 0 (0) | 3 (23) | 2 (15) | 5 (38) | 1 (8) | 2 (15) |
| Active anti-hormonal therapy |  |  |  |  |  |  |
| Yes | 0 (0) | 1 (13) | 2 (25) | 2 (25) | 2 (25) | 1 (13) |
| No | 0 (0) | 2 (17) | 0 (0) | 3 (25) | 4 (33) | 3 (25) |

| **Presence of Distant Metastases** |  |  |  |  |  |  |
| --- | --- | --- | --- | --- | --- | --- |
| Yes | 0 (0) | 0 (0) | 1 (50) | 0 (0) | 1 (50) | 0 (0) |
| No | 0 (0) | 3 (17) | 1 (6) | 5 (28) | 5 (28) | 4 (22) |

\*p=0.0018, \*\*p=0.0347, \*\*\*p=0.0238, \*\*\*\*p=0.0103, Fisher Exact, two-tailed. <sup>a</sup> N=87. One participant showed exercise-induced arrhythmia during pulse recovery, excluded from analyses. <sup>b</sup> N=24. No participants excluded. <sup>c</sup> N=20. Pulse recovery not determined for one participant, excluded from analyses. No.: Number.

### Specific Muscle Group Testing Revealed Strengths and Weaknesses Relevant to Exercise Prescriptions

Fifty-six percent of the cancer survivors presenting to clinic were offered abdominal and hip-flexor strength testing with a crunch test (Table 4). Overall 87% of those tested were able to complete the full 20 crunches in good form. The percentage completing 20 crunches ranged from 60% to 100% across the different step test groups. In contrast, successful completion rates of 20 modified push-ups from the knees were low across all groups and, in the end, only 21% were offered this test in clinic. The Squat Test was used as a means of more specific assessment of quadriceps, hamstring and gluteal muscle strength with 17% completing. Fifty-four percent of this proportion had an abnormal test. Ten percent of survivors were offered a plank test, which assessed transversus abdominis, rectus abdominis, internal oblique and external oblique muscles. The mean number of seconds held was 12+/-2 secs (mean +/ SEM) with no significant variations between step test groups. There were no significant associations between cardiorespiratory fitness testing and strength testing; however, performance of crunches, squat tests and planks enabled identification of those individuals within each group who required additional core muscle training prior to engaging in a vigorous cardiorespiratory regimen. So few individuals could perform a modified push-ups that this test was relatively non-informative for this population.

**Table 4:** Strength Testing Results By Step Test Category for Cancer Survivors Participating in Clinic.

|  | Step Test Category |  |  |  |  |  |  |
| --- | --- | --- | --- | --- | --- | --- | --- |
|  | Total Cohort | At Pace | Slower Self-Paced | Shorter Self-Paced | Flat Self-Paced | Contraindicated | Deferred |
| **Cancer Survivor Total, No. (%)** | 169 (100) | 88 (52) | 24 (14) | 21 (12) | 9 (5) | 19 (11) | 8 (5) |
| **Crunch Test, No. (%)** |  |  |  |  |  |  |  |
| Tested | 94 (56) | 55 (64) | 14 (58) | 15 (71) | 5 (56) | 3 (16) | 2 (25) |
| Not Tested | 75 (44) | 33 (36) | 10 (42) | 6 (29) | 4 (44) | 16 (84) | 6 (75) |
| No. Performed |  |  |  |  |  |  |  |
| 20 (maximal) | 82 (87) | 50 (90) | 12 (83) | 12 (82) | 3 (60) | 3 (100) | 2 (100) |
| 15-19 | 3 (3) | 1 (2) | 0 (0) | 1 (6) | 1 (20) | 0 (0) | 0 (0) |
| 10-14 | 3 (3) | 1 (2) | 2 (17) | 0 (0) | 0 (0) | 0 (0) | 0 (0) |
| 5-9 | 4 (5) | 3 (6) | 0 (0) | 1 (6) | 0 (0) | 0 (0) | 0 (0) |
| 0-4 | 2 (2) | 0 (0) | 0 (0) | 1 (6) | 1 (20) | 0 (0) | 0 (0) |
| **Push-Up On Knees, No. (%)** |  |  |  |  |  |  |  |
| Tested | 36 (21) | 20 (23) | 8 (33) | 5 (24) | 1 (11) | 0 (0) | 2 (25) |
| Not Tested | 133 (79) | 68 (77) | 16 (67) | 16 (76) | 8 (89) | 19 (100) | 6 (75) |
| No. Performed |  |  |  |  |  |  |  |
| 20 (maximal) | 11 (30) | 6 (30) | 3 (37) | 0 (0) | 1 (100) | 0 (0) | 1 (50) |
| 15-19 | 1 (3) | 1 (5) | 0 (0) | 0 (0) | 0 (0) | 0 (0) | 0 (0) |
| 10-14 | 2 (6) | 1 (5) | 0 (0) | 1 (20) | 0 (0) | 0 (0) | 0 (0) |
| 5-9 | 8 (22) | 6 (30) | 2 (26) | 0 (0) | 0 (0) | 0 (0) | 0 (0) |
| 1-4 | 14 (39) | 6 (30) | 3 (37) | 4 (80) | 0 (0) | 0 (0) | 1 (50) |
| **Squat Test, No. (%)** |  |  |  |  |  |  |  |
| Tested | 29 (17) | 18 (20) | 3 (12) | 4 (19) | 2 (22) | 2 (11) | 0 (0) |
| Not Tested | 140 (83) | 70 (80) | 21 (88) | 17 (81) | 7 (88) | 17 (89) | 8 (100) |
| Normal Result | 10 (44) | 7 (39) | 1 (33) | 0 (0) | 1 (50) | 1 (50) |  |
| Abnormal Result | 19 (66) | 11 (61) | 2 (67) | 4 (100) | 1 (50) | 1 (50) |  |
| **Plank, No. (%)** |  |  |  |  |  |  |  |
| Tested | 17 (10) | 8 (9) | 2 (8) | 5 (24) | 1 (11) | 1 (5) | 0 (0) |
| Not Tested | 152 (90) | 80 (91) | 22 (92) | 16 (76) | 8 (89) | 18 (95) | 8 (100) |
| Seconds Held Mean±SEM<br>(Range) | 12±2<br>(5-30) | 13±3<br>(5-30) | 10±0<br>(10) | 11±2<br>(5-20) | 15±0<br>(15) | 15±0<br>(15) |  |

No.: Number. SEM: Standard Error of the Mean

## DISCUSSION

This study demonstrates utility of a fitness and metabolism clinic for individualized fitness evaluation in cancer survivors. Modified Harvard Step Tests administered within the context of a comprehensive physical examination that included exercise history and targeted testing of specific muscle groups resulted in exercise prescriptions that moved survivors from sedentary to light exercise, light to moderate, or moderate to vigorous as appropriate. History and physical provided preliminary evaluation of fitness with identification of balance, musculoskeletal, deconditioning and frailty, and cardiac-related reasons to exclude step testing. However, not all of the reasons why cancer survivors were challenged by step test fitness testing were cancer related. Knee pain was the most common non-cancer related morbidity, reported by 11% of the cancer survivors presenting to clinic. Prevalence of knee pain and osteoarthritis has increased significantly in the US over past decades so it not unexpected that it was also found here [37,38].

In the general population fitness is positively associated with socioeconomic status and education [29]. Here all of the cancer survivors presenting to clinic had established residences, most were employed in economic sectors associated with a professional education and approximately two thirds lived with social support from family and/or life partners. However, similar to previous reports for cancer survivors, their current exercise activity was significantly less vigorous than their past activity and, only 38% (n=65) had an age-adjusted fitness rating from above average to excellent on the 3-minute, 96 +/-0 steps/min, 9 inch step test. Performance on the slower and shorter tests, while for purposes of stratification here using the full ranking range developed by the YMCA and reported as above average to excellent performance, does not actually reflect a high fitness rating as the tests themselves were significantly less vigorous. At the same time, the results showed that even these less vigorous tests stratified relative fitness, provided a baseline fitness level for later comparison, and the observed performance was critical for development of an appropriate exercise prescription to build cardiorespiratory fitness. A next step would be to develop appropriate age-adjusted fitness rankings for these alternative step tests, perhaps standardizing slower pace and duration, that would more accurately reflect fitness ranking. For example, it is possible the highest fitness ranking for these tests would be more appropriately referred to as average with performance then ranging from very poor-average for these tests. Although the flat test did not stratify into YMCA fitness ratings, it was useful for assessing balance and the first level of exercise, ability to walk.

It is recommended that cancer survivors receive medical clearance prior to initiating an exercise program. Here the majority of cancer survivors in general presented within two years of diagnosis, in general, an appropriate timeframe for intervention as the majority had completed surgical, radiation and/or chemotherapy. Hormonal therapy generally follows completion of these treatment modalities and extends for years therefore the relatively high proportion of cancer survivors on active hormone therapy (48%) was not unexpected in this population with a high rate of breast cancer. Different impacts of hormonal therapy on fitness have been described [11]. Here we found hormonal therapy positively associated with fitness ranking for the group completing the At Pace test but negatively associated for the Slower Self-Paced group and without association with the Shorter Self-Paced group. The uneven size of the groups may contribute to lack of a uniform finding but it is also possible that the significantly older ages of the Slower and Shorter Self-Paced groups as compared to the At Pace group plays a role as age is generally a fitness determinant. Another difference was the type of hormonal therapy. In the At Pace group 73% of those taking hormonal therapy were on tamoxifen while in the Slower Self-Paced group it was 55% and in the Shorter Self-Paced group it was 30%. In the future it may be useful to address possible interactions between age and/or type of hormonal therapy on fitness directly although this could be confounded by the restrictions on use of aromatase inhibitors in premenopausal women. A significant minority of presenting survivors (28%) were under active chemotherapy at the time of presentation. Chemotherapy was associated with lower fitness rankings for the At Pace group, similar to what has been previously reported, but not associated with the Slower Self-Paced, and associated with higher fitness rankings for the Shorter Self-Paced group, illustrating that even if survivors remain under chemotherapy they still can be effectively evaluated for an appropriately rigorous exercise prescription [39]. The same was true for survivors with distant metastases. With appropriate screening though H&P, 10/16 survivors with distant metastasis were able to participate in step testing, the majority (n=7) completed the At Pace test. Inclusion of survivors on active therapy and who may have distant metastases in fitness evaluation as appropriate is a current recommendation ACS recommendation [3].

For the At Pace group, BMI and initial pulse were both inversely associated with fitness. The lack of association with the Slower and Shorter Self-Paced tests may be due to their lower exertion levels. Age at time of testing and age at cancer diagnosis were both positively correlated with the ability to complete the 3-minute, 96 +/-0 steps/min, 9 inch step test as was both past and current vigorous exercise histories. All of these factors are also correlated with higher fitness levels in the general population, illustrating that while cancer survivors may have the extra burden of recovery from surgery and radiation and possible on-going chemotherapy and hormone therapy, in general, their behavior is very similar to the general population [29,40].

The approach utilized represents a cost-effective model for addressing exercise prescription in survivorship care. Evaluation was incorporated into a primary care comprehensive physical examination appropriate for cancer survivors as they transition away from the acute management of their disease. The set-up is consistent with recent AHA recommendations for health assessments in the youth population and beneficial for all stakeholders [36]. For health insurers, it effectively incorporates both short-term and long-term evaluation of cancer survivors into a standard visit. It may also improve utilization of wellness initiatives promoted by various insurers. For instance, Medicare Advantage plans covering fitness memberships have found to both attract and keep a healthier proportion of the population using Medicare [41]. For patients, provision of fitness testing in the office enabled a hands-on experience in a safe environment. If a patient had issues in balance, asymmetry, and perceived exertion in step testing, the provider was able to intervene and demonstrate the correct form. Inclusion of specific muscle group testing enabled the provider to directly assess core strength required for initiating effective cardiorespiratory exercise and correct form as needed. Crunches and squats were the most widely applied test, which were able to identify the minority of survivors who required attention to abdominal core musculature and the more significant percentage of survivors who would benefit from exercises to improve lower extremity strength. Although this study did not examine prospective adherence, it is known that in-office demonstration of other tasks, like insulin injection and inhaler use, increases patient adherence [42,43]. Furthermore, research suggests that if prescribed exercise is too intense, patients are more likely to drop out [44]. The individualized exercise prescription ensures that patients begin with the correct level of exertion to increase both program satisfaction and adherence.

Limitations of our study included retrospective chart review design, the focus on a higher socioeconomic population, and over-representation of female, breast cancer survivors. However, it does illustrate the approach of incorporating step testing and focused muscle strength testing into a comprehensive physical exam for fitness evaluation and exercise prescription for cancer survivors. Further prospective research is needed to assess patient adherence and fitness progression following exercise prescription with expansion into populations with different socioeconomic status who may lack social support and/or a permanent residence. The use of telemedicine for follow-up visits may also be a target for future investigation, in an age where virtual engagement is increasingly utilized in health care, particularly for healthy lifestyle promotion [45].

## Data Availability

All data referred to in the manuscript is contained either within the Tables included in the manuscript or the supplementary figure file.

## Acknowledgements

The authors thank the Lombardi Survivorship Research Initiative for advice and encouragement and Georgetown Lombardi Comprehensive Cancer Center and MedStar Georgetown University Hospital staff for their assistance and guidance.

